# Endemicity is not a victory: the unmitigated downside risks of widespread SARS-CoV-2 transmission

**DOI:** 10.1101/2022.03.29.22273146

**Authors:** Madison Stoddard, Alexander Novokhodko, Sharanya Sarkar, Debra Van Egeren, Laura F. White, Natasha S. Hochberg, Michael Rogers, Bruce Zetter, Diane Joseph-McCarthy, Arijit Chakravarty

## Abstract

We have entered a new phase of the ongoing COVID-19 pandemic, as the strategy of relying solely on the current SARS-CoV-2 vaccines to bring the pandemic to an end has become infeasible. In response, public-health authorities in many countries have advocated for a strategy of using the vaccines to limit morbidity and mortality while permitting unchecked SARS-CoV-2 spread (“learning to live with the disease”). The feasibility of this strategy is critically dependent on the infection fatality rate (IFR) of COVID-19. An expectation exists, both in the lay public and in the scientific community, that future waves of the virus will exhibit decreased IFR, either due to viral attenuation or the progressive buildup of immunity. In this work, we examine the basis for that expectation, assessing the impact of virulence on transmission. Our findings suggest that large increases in virulence for SARS-CoV-2 would result in minimal loss of transmission, implying that the IFR may be free to increase or decrease under neutral evolutionary drift. We further examine the effect of changes in the IFR on the steady-state death toll under conditions of endemic COVID-19. Our modeling suggests that endemic SARS-CoV-2 implies vast transmission resulting in yearly US COVID-19 death tolls numbering in the hundreds of thousands under many plausible scenarios, with even modest increases in the IFR leading to an unsustainable mortality burden. Our findings thus highlight the critical importance of enacting a concerted strategy (involving for example global access to vaccines, therapeutics, prophylactics and nonpharmaceutical interventions) to suppress SARS-CoV-2 transmission, thereby reducing the risk of catastrophic outcomes. Our findings also highlight the importance of continued investment in novel biomedical interventions to prevent viral transmission.

## Introduction

As the COVID-19 pandemic continues unabated, it is easy to forget that the consensus belief not so long ago (both in the scientific community ^1–3^ and the popular press ^4–7^) was that the deployment of SARS-CoV-2 vaccines would bring the pandemic to an end.

Waning immunity ^8,9^ and the rapid evolution of viral immune evasion ^10,11^ have limited the ability of vaccines to block transmission ^12,13^ and dimmed the prospects for herd immunity to SARS-CoV-2. Of the 16 countries in the world with two-dose vaccination rates above 70% of the population, 12 have experienced their highest level – and the other 4 their second-highest level – of disease transmission during the omicron wave (see Supplementary Material S2). Further underscoring the infeasibility of using the existing vaccines alone to eliminate SARS-CoV-2, settings with extremely high vaccination rates have seen large chains of transmission, even in the presence of other mitigation measures ^14^ and super-spreader events have been demonstrated in some cases to be driven by vaccine breakthrough cases ^15^.

With complete eradication of SARS-CoV-2 (the viral pathogen responsible for COVID-19) seemingly off the table, some public health authorities ^16–18^ now advocate for a strategy of “learning to live with the virus”. This transition from “pandemic” to “endemic” conditions is thought to be possible as the rate of viral transmission is eventually maintained at a steady-state level by the limited availability of susceptible hosts. In practice, this strategy emphasizes relying on the vaccines’ high level of protection against severe acute disease and hospitalization to limit short-term morbidity and mortality, without taking other steps to limit transmission. A critical assumption underpinning this public health strategy is that infections with SARS-CoV-2 will lead to milder outcomes over time, either due to the progressive buildup of immunity within individuals or due to viral attenuation.

The progressive buildup of immunity hypothesis posits that upon repeated infections (or vaccinations), individuals develop increased immunity to SARS-CoV-2 infections, which in turn leads to a reduced risk of death upon SARS-CoV-2 infection. This hypothesis relies on the level of immunity increasing within the population. However, the broad population heterogeneity of rates of decline of immunity, coupled with viral immune evasion, may make it challenging for populations to build immunity to SARS-CoV-2 over time. Consistent with this, Bayesian modeling based on Census Bureau data (accounting for waning vaccinal immunity, immune evasion and the pace of vaccinations) suggests that the effective protection against infection in December 2021 (21%, against the omicron variant) was lower than the effective protection against infection in January 2021 (25%, against the ancestral strain)^19^.

The viral attenuation hypothesis, on the other hand, posits that natural selection will favor viral variants with reduced virulence, leading to an evolutionary ratchet that monotonically reduces the mortality burden of SARS-CoV-2 infections over time. The strong phrasing of this hypothesis (“emergent viruses evolve to become less virulent over time”) is a commonly held belief ^20,21^ that is demonstrably false ^22^. While in some settings there may be a tradeoff between virulence and transmission such that increased virulence leads to reduced transmission, this is not a general rule. A number of emergent viruses in other species have evolved to be both more transmissible and more virulent over time: examples include feline calicivirus ^23^, myxomatosis in rabbits ^24–26^ and H5N2 influenza in birds ^27,28^. The historical record also contains multiple examples of human pathogens whose virulence has increased over time. HIV virulence has been shown to have increased steadily since its emergence ^29,30^, underscored by the recent discovery of a highly virulent strain of HIV that has been circulating for several years ^31^. The second wave of the 1918-1919 influenza pandemic was substantially more deadly ^32^ than the first, with a change in the impacted population such that younger individuals had an elevated risk of death. This increased virulence is attributable to viral evolution, as experiments in *in vitro* and animal systems suggested that the coordinated expression of eight genes unique to the 1918 virus was responsible for the increased lethality ^33,34^. For other human pathogens, such as smallpox, virulence fluctuated wildly from one wave to the next ^35,36^, with the high-virulence (“variola major”) strains showing functional differences from the low virulence (“variola minor”) strains ^37,38^ in *in vitro* and animal studies. The instability of smallpox virulence over time contradicts the notion of obligatory viral attenuation and may foreshadow similar behavior during the current pandemic. For SARS-CoV-2, the infection fatality rates (IFR) for the alpha, beta, gamma, and delta variants of concern (VOCs) were higher than that of the ancestral strain, while the IFR of omicron appears substantially lower than that of the ancestral strain (see Supplementary Table S3).

In addition to changes in the intrinsic virulence of the virus, changes in medical practices or patient characteristics can also lead to substantial shifts in the IFR for SARS-CoV-2. Hospital capacity, treatment protocols, availability and effectiveness of therapies, population age structure ^39^, pollution exposure ^40^, seroprevalence ^41^, and numerous other factors have been shown to impact the IFR (See Supplementary Table S1 for more detail).

Thus, the expectation of a monotonic reduction in SARS-CoV-2 IFR over time deserves closer examination. With this in mind, we sought to explore the effect of increased virulence on the ability of SARS-CoV-2 to transmit efficiently and to predict the impact of changes in IFR on the practicality of “learning to live with the disease”. Using a range of plausible reinfection fatality rates and durations of sterilizing immunity, we explored the effects of an endemic or hyperendemic SARS-CoV-2 virus on yearly US COVID-19 mortality.

## Results

### Loss of viral fitness incurred by patient death is minimal

Figure 1a overlays the transmission probability distribution function (PDF) ^42^, which describes the distribution of transmissibility over time during one individual’s SARS-CoV-2 infection, and the fatal outcome PDF ^43^, which describes the likelihood of death over time since symptom onset in fatal cases. The time to death from COVID-19 is substantially longer than the time to transmission for the virus. The average loss of transmissibility due to a fatal outcome is determined by multiplying a patient’s probability of having died over time (the CDF of fatal outcome by time post onset of symptoms) by the transmissibility over time (detailed calculation is provided in the Methods section). Based on these distributions, we determined that the expected loss of infectivity incurred when a patient dies is approximately 1.3% of that patient’s overall propensity to transmit (Figure 1b). Thus, the overall loss of transmissibility for a lethal SARS-CoV-2 strain is 1.3%*IFR, assuming the PDFs for fatal outcome and infectivity over time are unchanged relative to the ancestral strain. Alarmingly, a strain that proved lethal in 100% of patients would thus only suffer a 1.3% loss of transmissibility, as nearly 99% of transmission precedes death in fatal cases. We note that this 1.3% loss of transmissibility is much smaller than previously observed increases in transmissibility accompanying new variants (Supplementary Table S3), suggesting that a minor loss in transmissibility due to a higher IFR could be readily overcome by improvements in transmissibility ^44,45^. Since changes in the IFR do not significantly impact the transmissibility of SARS-CoV-2, there is no evolutionary pressure favoring a reduction in COVID-19 disease severity. This implies that the intrinsic IFR (the IFR in the absence of novel interventions and in an immunologically naïve population) is not likely to steadily decrease over time.

**Figure 1.**
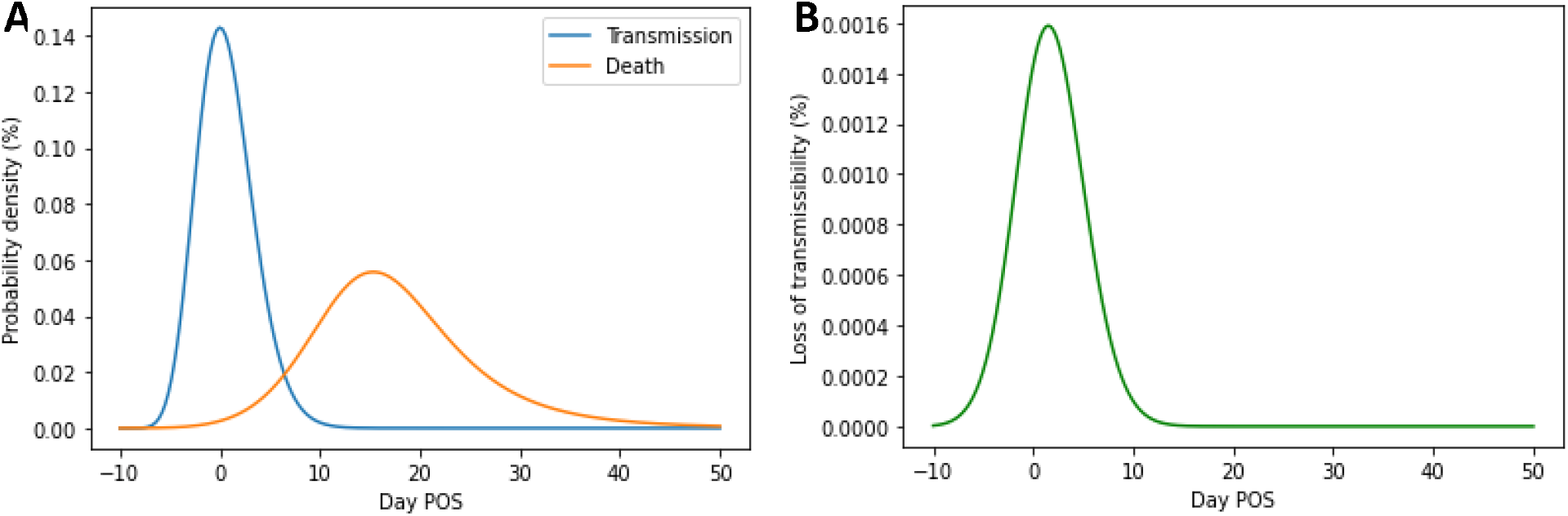
Transmission as a function of time post onset of symptoms (POS) is minimally impacted by fatal COVID-19 outcomes. A) PDFs for COVID-19 death and transmission over time. B) Loss of fractional transmissibility over time in fatal cases.

We note that it is possible for novel variants to emerge that have a faster timeline for death such that their transmission will be reduced. However, by definition, such variants will be placed at an evolutionary disadvantage, and are not relevant to the analysis here. In this context, the trajectory of SARS-CoV-2 evolution so far reveals that an increased IFR is not obligatorily associated with a faster timeline to death or reduced transmission. For example, the delta variant (around 80% more transmissible) also had around a 50% higher risk of death (see Supplementary Table S3 for more details). The wide spacing between the time of peak transmissibility and the time of peak mortality risk suggests that the virus may be able to continue to increase transmissibility and virulence at the same time.

### Steady-state transmission of SARS-CoV-2 is extensive

To determine the consequences of permitting widespread SARS-CoV-2 transmission, we simulated the endemic spread of SARS-CoV-2 using a susceptible-exposed-infected-recovered-susceptible (SEIRS) model accounting for waning of natural immunity against reinfection (Methods). Endemic disease spread is characterized by a steady rate of reinfections required to maintain the steady-state level of immunity under conditions of immune waning. In Figure 2, we explored the model-predicted steady-state level of US SARS-CoV-2 transmission under six vaccination scenarios: 0%, 50%, or 90% reduction in risk of infection (VEi) with 70% or 100% vaccine uptake in the population. These simulations demonstrate the challenge inherent in managing a highly transmissible, endemic disease conferring only short-term immunity: extreme levels of infection. Under optimistic assumptions – 70% immunization with a vaccine that reduces risk of infection by 90%, an intrinsic reproductive number (R_0_) of 5 and 18-month duration of natural immunity – over 50 million US infections can be expected annually. An increase in transmissibility to an R_0_ of 8 results in nearly 100 million infections, and persistent immune evasion resulting in a 9-month duration of natural immunity (with R_0_ = 5) would also increase infection counts to approximately 100 million. An accompanying drop in vaccine protection against infection to 50% would result in a staggering infection burden approaching 300 million US infections annually. However, complete suppression of SARS-CoV-2 spread is possible with a vaccine that is highly effective against infection and widely accepted by the population (Figures 2c and 2f).

**Figure 2.**
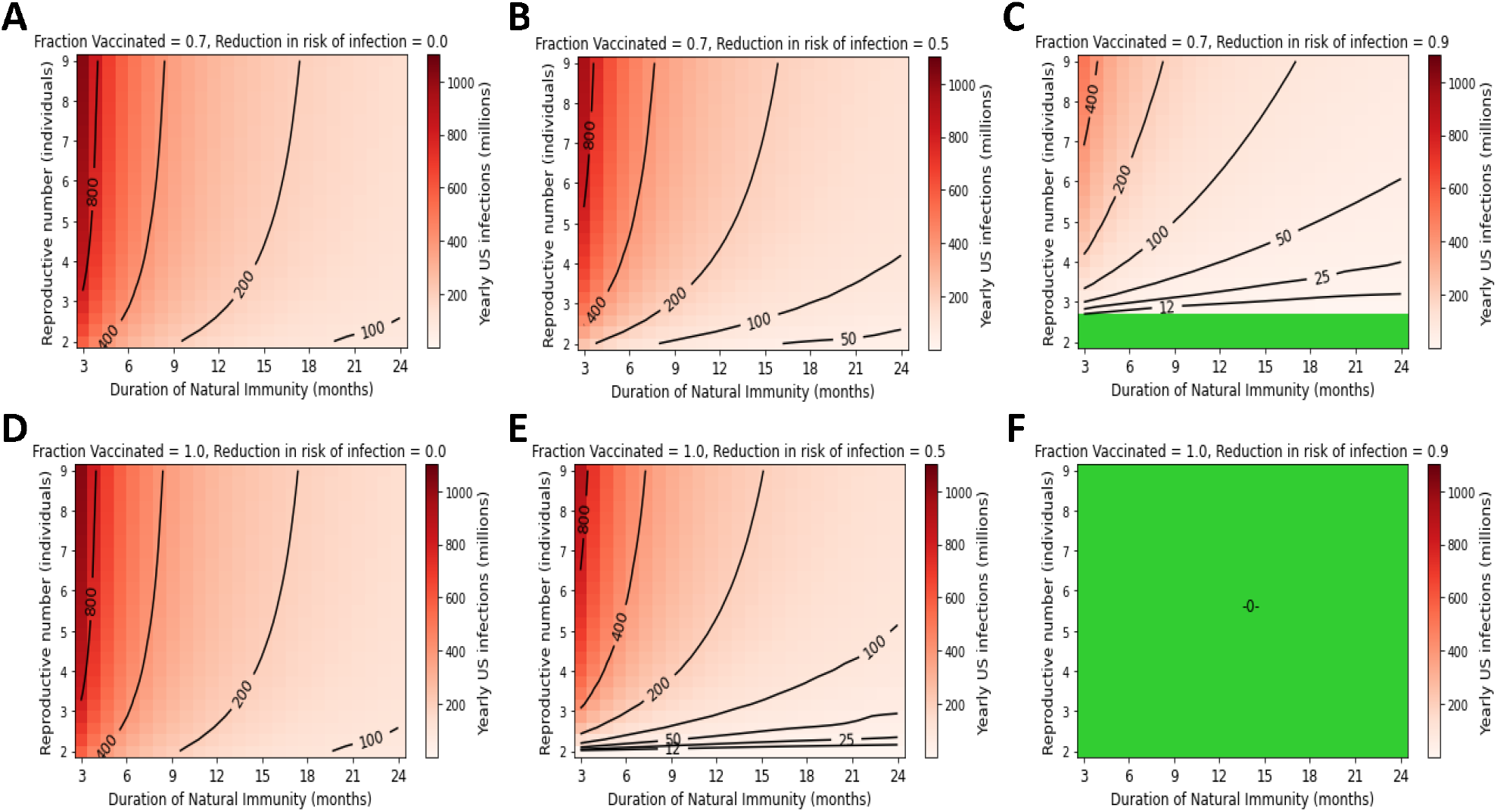
High yearly US infection counts persist under endemic conditions without vaccines that prevent transmission. Yearly US SARS-CoV-2 infections under the following conditions for vaccine compliance and VEi: A) 70% vaccinated with 0% VEi, B) 70% vaccinated with 50% VEi, C) 70% vaccinated with a 90% VEi, D) 100% compliance with a vaccine with 0% VEi, E) 100% compliance with 50% VEi, F) 100% compliance with 90% VEi. Green regions represent complete suppression of SARS-CoV-2 transmission.

### Endemic SARS-CoV-2 death tolls are highly sensitive to changes in IFR and duration of immunity

For SARS-CoV-2, the infection fatality rate (IFR) has diverged considerably from the 0.7% of the ancestral strain ^46^, as IFRs for the VOCs have ranged from 0.21% for omicron to 1.58% for delta (see Supplementary Table S3). As the delta variant was not directly descended from any of the preceding VOCs, and omicron was not directly descended from delta ^47^, IFRs ranging from 0.2% to 1.6% can be considered as the baseline for SARS-CoV-2. In addition, IFRs of up to 3% have been observed at various points during the pandemic, due to changes in local conditions (See Supplemental Table S1). In this study we have considered IFRs between 0.05% and 5%, with 0.7% considered the baseline, corresponding to the ancestral strain ^46^.

In Figures 3 and 4, we explored the sensitivity of annual US COVID-19 fatalities to IFR, R_0_, and duration of natural immunity under 70% and 100% vaccine acceptance, respectively. We explored multiple R_0_ and VEi conditions but assumed vaccine efficacy against mortality given infection (VEm) is 90% under all scenarios. In these plots, yellow shading represents regions of parameter space where US COVID-19 deaths are predicted to exceed the approximately 650,000 yearly deaths from heart disease, the current leading cause of death^48^. The black point represents best estimates for the IFR and duration of natural immunity for the ancestral strain. We note that under conditions of an R_0_ of 5, 70% vaccination under a VEi of 50%, and best estimate parameters for IFR and natural immunity, the model predicts approximately 420,000 US COVID-19 deaths annually. For reference, this is comparable to the approximately 460,000 COVID-19 deaths reported by the CDC for 2021 ^49^.

**Figure 3.**
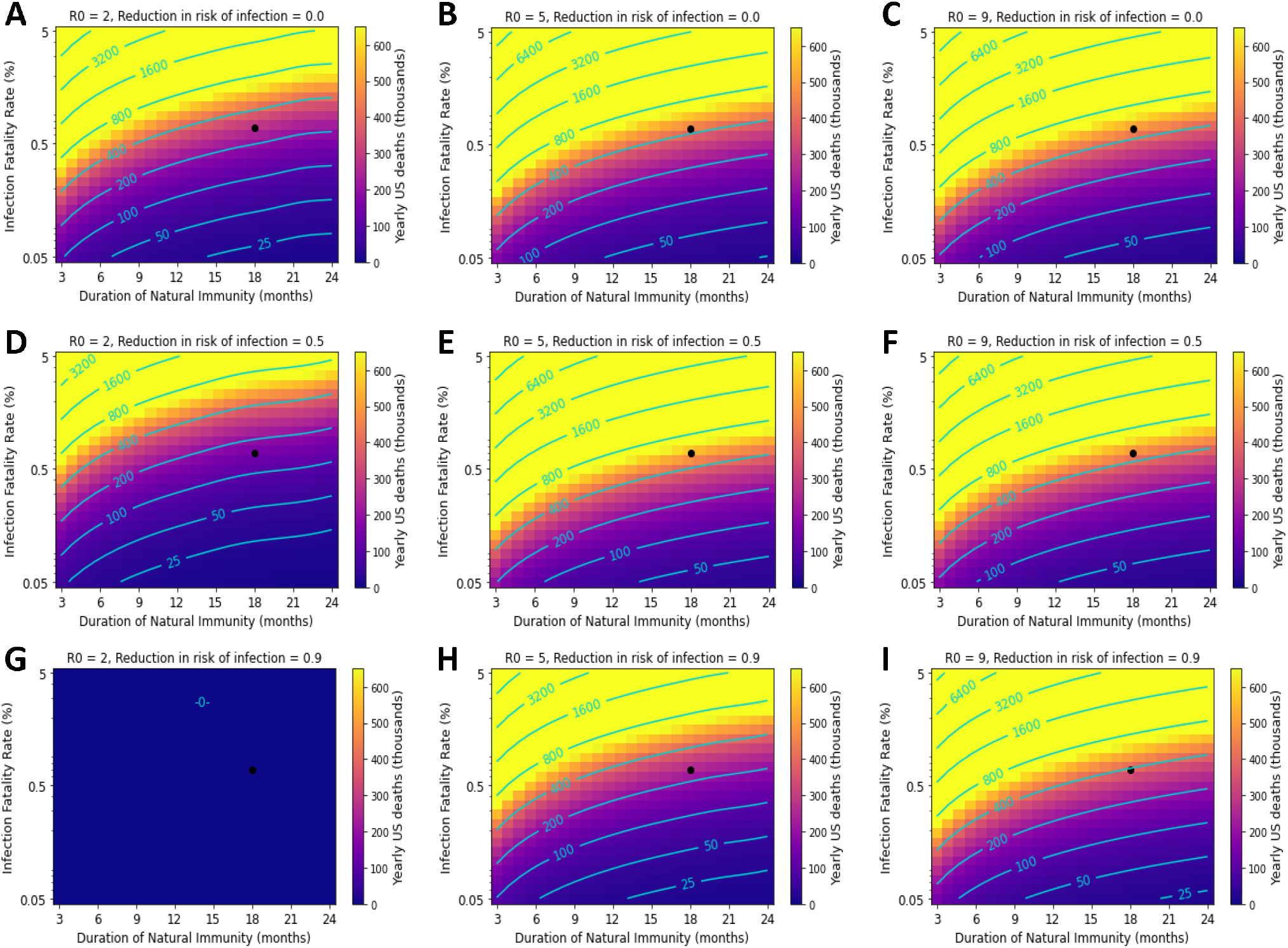
Variation in the duration of natural immunity and IFR can result in catastrophic death tolls. The black point represents parameter values corresponding to best-estimates of immunity and IFR for ancestral SARS-CoV-2. Yearly US COVID-19 deaths under the following transmissibility (R_0_) and VEi conditions: A-C) 0% VEi and R_0_ of 2, 5, and 9; D-F) 50% VEi and R_0_ of 2, 5, and 9. G-I) 90% VEi and R_0_ of 2, 5, and 9. Vaccine compliance is 70% and VEm is 90% in all panels.

**Figure 4.**
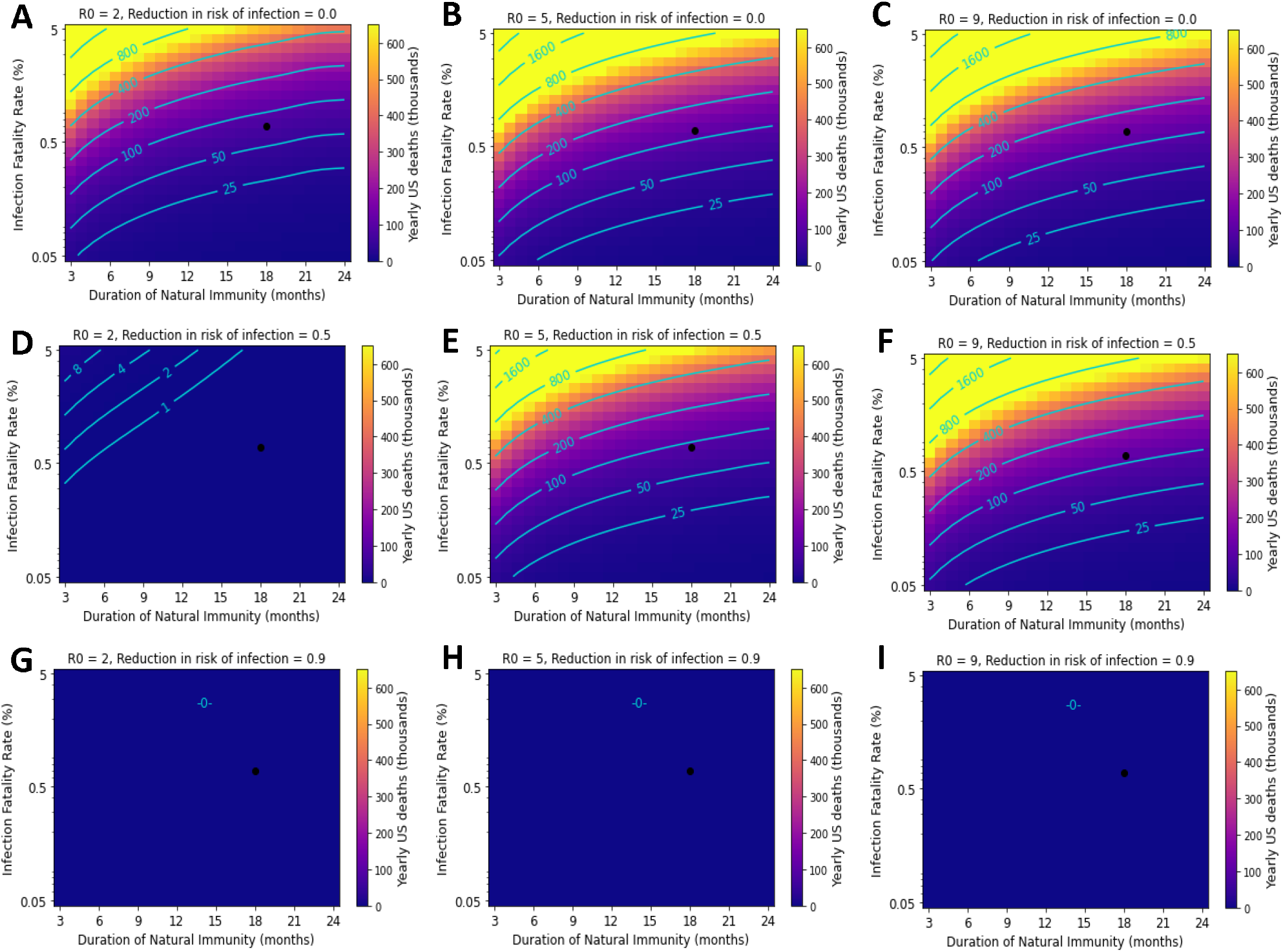
SARS-CoV-2 containment through highly effective vaccination mitigates mortality risks related to variability in IFR and immunity. Black point represents parameter values corresponding to best-estimates of immunity and IFR for ancestral SARS-CoV-2. Yearly US COVID-19 deaths under the following transmissibility (R_0_) and VEi conditions: A-C) 0% VEi and R_0_ of 2, 5, and 9; D-F) 50% VEi and R_0_ of 2, 5, and 9. G-I) 90% VEi and R_0_ of 2, 5, and 9. Vaccine compliance is 100% and VEm is 90% in all panels.

Our model predicts that COVID-19 is likely to become the leading cause of death in the US under many scenarios. For example, for a hypothetical SARS-CoV-2 variant with an R_0_ of 5, an IFR of 1%, and a 12-month duration of natural immunity, approximately 700,000 US COVID-19 deaths could be expected per year if a vaccine preventing 90% of infections were administered to 70% of the population. We observe that despite a high degree of vaccine efficacy – 90% reduction in risk of infection and 90% reduction in risk of death given infection– the region in which US COVID-19 deaths under endemic conditions rival influenza deaths (12,000 – 52,000 per year, according to the CDC^50^) is small and would require a significant reduction in IFR.

In Figure 4, we performed the same parameter sweep under the assumption that the entire population is vaccinated. Under these conditions, a much more favorable outcome is apparent in some scenarios: complete suppression of SARS-CoV-2 (Figures 4g-i), resulting in essentially zero yearly deaths (Figure 2). This is possible when a sufficiently high proportion of the population is vaccinated with a vaccine that prevents most infection. Although the near-suppression scenario (Figure 4d) is favorable, the failed suppression scenarios (Figures 4a-c and 4e-f) entail large death annual COVID-19 tolls under most conditions despite 100% acceptance of a vaccine that prevents 90% of fatalities in breakthrough cases. As shown in Figure 2, transmission under these scenarios is simply too high for population-level mortality to be controlled by such a vaccine. Additionally, we note that the COVID-19 death counts show a linear dependence on changes in IFR.

Mortality under these high-transmission steady-state scenarios is also impacted by changes in the durability of natural immunity, while changes in R_0_ have relatively little impact. This is because R_0_ determines the level of immunity required to maintain the steady-state according to a saturating relationship (1-1/R_0_), while the durability of natural immunity determines the rate at which immunity must be replenished to attain this steady-state level. The IFR describes the direct proportionality between the number of SARS-CoV-2 infections and COVID-19 fatalities.

In Supplementary Figures 1 and 2, we explore the consequences of SARS-CoV-2 endemicity under a vaccine with a VEm of 70% (compared to 90% in Figures 3 and 4). Based on these plots, we conclude that small losses in vaccine efficacy against mortality can result in substantial increases of population-level mortality. For example, the model prediction for COVID-19 mortality under best-estimate parameters (an R_0_ of 5, 18-month duration of natural immunity, and 70% coverage with a vaccine with 50% VEi) is approximately 400,000 if the vaccine prevents 90% of mortality after infection. If this vaccine’s VEm reduces to 70% (Figure S1), the predicted death toll rises by 50%, approaching 600,000 per year.

### Relationship between R_0_ and yearly death toll is saturating

Figure 5 elucidates the relationship between R_0_ and yearly US COVID-19 mortality. As R_0_ increases, the yearly endemic death toll increases, but this relationship saturates as R_0_ increases. This means that in scenarios where SARS-CoV-2 is contained by a slim margin, significant outbreaks may be possible with small increases in R_0_ or losses of VEi. Additionally, changes in contact behavior or vaccine efficacy against infection are most impactful when the R_0_ is closer to 1, while measures that minimally reduce transmission under a high R_0_ may have little impact on overall mortality. Under endemic conditions, immune evasion resulting in changes in vaccine efficacy or the durability of natural immunity are likely to be more impactful than further increases in transmissibility.

**Figure 5.**
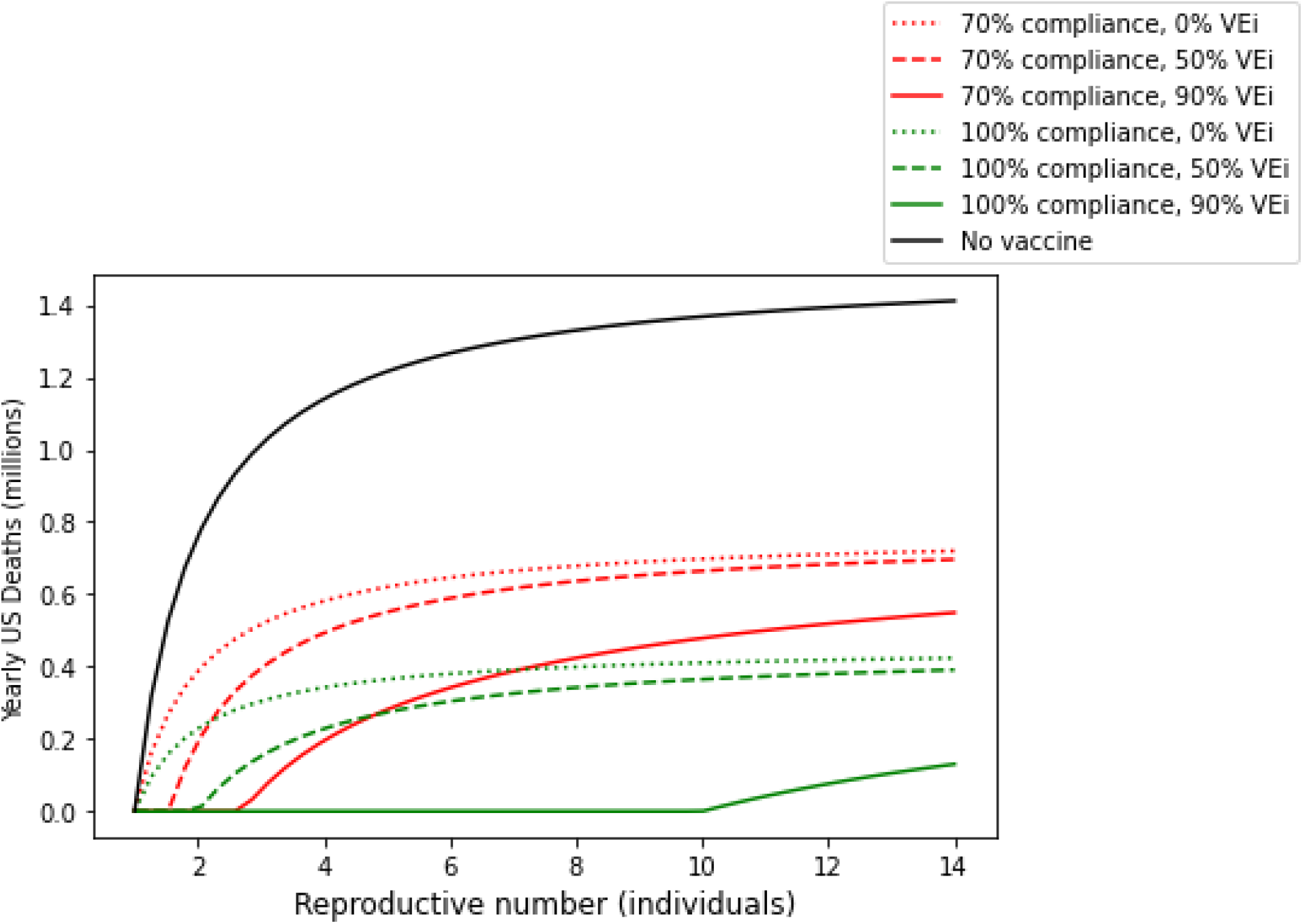
Relationship between R_0_ and yearly US deaths is nonlinear. Assuming the duration of natural immunity is 18 months, the IFR is 0.7%, and VEm is 90%, endemic US COVID-19 death tolls were simulated under a variety of vaccination scenarios.

### Emergence of new variants may rapidly drive infection levels exceeding the steady state

The emergence of new SARS-CoV-2 variants with increased transmissibility and/or immune evasion may result in significant infections above the steady-state level in short periods of time, as observed during the omicron wave. For example, the recent omicron wave (12/8/2021-2/24/2022) led to 30 million recorded COVID-19 cases ^51^, which corresponds to at least 75 million infections using an infection: case ratio of 2.5 ^52^. (This is the low end of the range of published estimates ^53–55^ and likely to be an undercount due to the high positivity rate ^56^ and reduced sensitivity to detection ^57^ seen during the omicron wave). Table 1 illustrates the challenges with managing death tolls from waves of this size: precedented shifts in IFR (such as those described in Supplemental Table S1) can result in mass casualties in short periods of time.

**Table 1.** Large waves of infection can lead to catastrophic death tolls with IFRs that are consistent with previous variants.

| **Infections \ IFR** | **0.5%** | **1%** | **2%** |
| --- | --- | --- | --- |
| **50 million** | 250,000 | 500,000 | 1,000,000 |
| **100 million** | 500,000 | 1,000,000 | 2,000,000 |
| **200 million** | 1,000,000 | 2,000,000 | 4,000,000 |

## Discussion

The work in this paper demonstrates the risks of COVID-19 management strategies that focus on limiting disease severity while permitting unmitigated spread. The high level of endemic disease propagation will prove challenging for healthcare systems to manage effectively, jeopardizing the ability of healthcare professionals to detect disease when it is most tractable to antivirals, identify patients at risk of severe outcomes, and optimally distribute treatment. This vast infection burden can be expected to translate into hundreds of thousands of COVID-19 fatalities even if vaccines reduce the risk of COVID-19 mortality by 90% or more. Additionally, these steady-state, endemic disease conditions may be interrupted by waves of transmission driven by immune-evading variants such as omicron. Most concerningly, SARS-CoV-2 may not be subject to evolutionary pressure favoring a lower virulence rate, and slight changes in the IFR of novel variants may lead to unanticipated – and potentially catastrophic – public health outcomes on both a chronic and an acute basis.

Overly optimistic predictions about the end of the ongoing pandemic ^1–7^ have tremendously complicated the public-health response to the crisis. Two aspects of viral behavior in particular were underestimated for SARS-CoV-2: its evolutionary potential, and the challenges inherent in a vaccine-only strategy (due to waning vaccinal immunity and low population-level compliance). These risks were in fact predictable. The impact of waning vaccinal immunity was identified as a threat to the feasibility of achieving vaccine-mediated herd immunity well before it came to pass ^58,59^. The rapid emergence of immune evasion in response to widespread population immunity was predicted by us and others ^10,60^, as was the infeasibility of relying on vaccines alone to permit a return to pre-pandemic conditions ^58,61,62^. Going forward, risk mitigation for this pandemic is threatened by an insufficient examination of the full downside potential of the situation at hand. In particular, the prediction of sustained low IFRs for future SARS-CoV-2 variants is an optimistic one. We show in this paper that it lacks a rigorous theoretic justification, and the consequences of this miscalculation could be immense.

The public (and public-health authorities) have taken “learning to live with this disease” as an inevitable consequence of the ineffectiveness of measures to reduce widespread transmission. This frames a false dichotomy ^63^ between eliminating SARS-CoV-2 and permitting its rampant spread. While it is relatively unrealistic to eliminate SARS-CoV-2 in the short term, reducing transmission is a necessary first step in managing the public-health burden of this disease. Many other pathogens that are considered to be extremely difficult to eliminate (such as influenza, tuberculosis, and malaria) have been the subject of long-term, globally-coordinated efforts at disease suppression. Accepting that tuberculosis is difficult to eliminate, for example, is not synonymous with encouraging its unrestrained spread across the globe.

Using the United States as an example, we note that the COVID-19 fatalities associated with the ancestral variant’s IFR (0.7%), vaccine parameters, and estimated duration of sterilizing immunity for COVID-19 (18 months) can be estimated at around 450,000 per year. (For reference, there were 460,000 deaths due to COVID-19 in the US during 2021^49^, and the “mild” omicron wave caused 150,000 deaths in the span of two months^49^.) The death toll due to COVID-19 vastly exceeds the mortality burden of the other leading infectious diseases in the US^64^. Accepting a new leading cause of death in the United States for the indefinite future will have profound impacts on life expectancy (as estimated by others based on 2020 data ^65,66^), and public-health planning should treat these impacts as the best-case scenario, instead of optimistically planning for them to reduce over time.

In fact, our work suggests that “learning to live with the disease” leads to a fragile outcome where the morbidity and mortality burden of the pandemic can be dramatically impacted by small shifts in IFR. The global experience with the omicron variant demonstrates that new waves of disease driven by emergent SARS-CoV-2 variants can spread extremely quickly, building to high levels of disease before the IFR can be reliably estimated. In the case of omicron, preliminary estimates suggest that the IFR is considerably lower than that of the ancestral strain; this work suggests that such an outcome is not guaranteed in the future. In a future scenario where an omicron-like variant sweeps quickly through the global population, but this time with a catastrophically high IFR, the unanticipated, lagging wave of death will be difficult to avoid after the fact of widespread infection. Our work also shows that the IFR need not be that high to cause catastrophe: an IFR of 1% is within the range of observed SARS-CoV-2 IFRs (see Supplementary Table S3) and would result in vast, rapid loss of life under an omicron-like variant wave. A significant public health risk at this point is the emergence and rapid spread of a new variant with an unexpectedly high IFR that only becomes apparent after it is too late to mitigate transmission.

Several authors have taken the position that, while increases in viral virulence may be possible, they are not likely. Using the analogy of wearing seatbelts when in a car, a worst-case scenario does not have to be likely for the risk to be worth mitigating. Increases in the IFR can occur due to direct virological factors (changes in viral load or immune evasion), or due to indirect factors (such as changes in viral tropism or pathology that erode the effectiveness of ICU interventions currently suppressing the IFR). Small, precedented changes in IFR (such as the changes described in Supplementary Table S1, for example) could lead to significant increases in COVID-19 death tolls in the US.

From the standpoint of evolution, there are at least two mechanisms by which a virus can simultaneously access higher lethality and improved transmission - increased viral load and innate immune evasion. Increased viral loads have been demonstrated to improve transmissibility as well as increase virulence for other diseases (^23, 24–26 27,28, 31^). The alpha and delta variants of SARS-CoV-2 were associated with increased viral load ^67–69^ as well as increased virulence (^70, 71^) relative to the ancestral strain. Innate immune suppression has been associated (in the case of other viruses) with improved reinfection potential, as well as dramatic increases in mortality (for example with the rabbit disease myxomatosis ^72^). SARS-CoV-2 is proficient at suppressing the innate immune response ^73–75^, targeting key innate immune effectors such as Type I interferon signaling ^76,77^ to delay the emergence of symptoms until after transmission has peaked. Many SARS-CoV-2 viral proteins have been implicated in innate immune suppression, such as ORF9b ^78,79^, ORF9c ^80,81^, Nsp1 ^82,83^, N protein ^84,85^, ORF3b ^85^, ORF6 ^84,85^, and ORF8 ^84^. This trait is not mediated by the spike protein (which dominates the antibody-mediated immune response ^86,87^, and thus can be expected to evolve independently of immunogenicity. Notably, enhanced innate immune evasion has already been observed for SARS-CoV-2, as recent variants of SARS-CoV-2 (alpha, delta, and omicron) all demonstrate robust overexpression of the N protein, as well as the protein products of the Orf 9b and Orf 6 genes ^88^. The unique aspects of SARS-CoV-2 transmission thus provide a biological basis by which increased virulence may provide a fitness advantage to future variants of SARS-CoV-2.

Our work has several key limitations. The model linking reinfection frequency, IFR and death toll is not age stratified, so it does not account for greater vaccine uptake in older populations. As a result, the model may overstate the death tolls, as risk of death is strongly age-dependent in COVID-19 ^89^. Offsetting this limitation is the finding that the benefit of vaccinal immunity appears to be age-dependent ^90^, and so the higher vaccine uptake in older populations may be undermined by a lower level of vaccinal efficacy over time. Additionally, the model does not account for evolution-mediated vaccine resistance or waning of vaccinal immunity and thus assumes vaccines retain their high efficacy over the simulation interval. This is likely to be an optimistic assumption and will also have the impact of mitigating death tolls. Our work does not explicitly model vaccines or boosters-a full exploration of the impact of vaccines on viral evolution is outside the scope of this work but explored by us elsewhere (manuscript in preparation). Similarly, the interplay between the kinetics of antibody decay and population heterogeneity in the rate of waning of natural and vaccinal immunity will impact the level of protection that vaccines provide, and a full treatment of these effects is outside the scope of this work but described in a different work by us (manuscript in preparation). As is true of all SEIR-type models, ours assumes homogenous population mixing and thereby overestimates the kinetics of viral spread and can underestimate the benefits of reduction in transmission at low viral prevalence. However, given the long time-frame of our model, these kinetics are not expected to substantially impact our conclusions. Although the model predicts long-term outcomes under endemic conditions, it is not designed to account for the impact of changes in population size over time due to excess COVID-19 mortality. We explore IFRs up to 5%, but the true span of possible IFRs may be larger (for example, case fatality rate estimates for coronaviruses SARS and MERS-CoV span ranges between 10-50% ^91–93^ and 20-40% ^94–96^, respectively). Lastly, our model assumes that natural immunity does not provide protection from mortality beyond protection from infection. The impact of a durable shift in IFR for reinfections can be estimated by selecting a lower IFR estimate in the analyses provided to reflect endemic conditions (under which virtually all infections will be reinfections, apart from those in young children). The work presented here should be viewed not as a specific prediction about the future, but rather as an exploration of the strategic implications of permitting widespread viral transmission while relying on vaccines to limit short-term morbidity and mortality.

In this context, given the poor performance of the first generation of SARS-CoV-2 vaccines in limiting variant spread, our work points to the critical importance of reinvesting in biomedical interventions for this disease. Designing biomedical interventions (antiviral prophylactics, mucosal vaccines) that can reduce transmission while resisting viral immune evasion remain a crucial unmet need in the current crisis. Antiviral prophylactics can provide vital assistance to vaccines by providing an orthogonal selection pressure (on non-spike proteins) that retards the emergence of novel immune-evasive variants. Multiple groups have reported the robust induction of mucosal immunity with nasal SARS-CoV-2 vaccines ^97–105^, and designing such vaccines to be robust to viral evolution would provide a powerful tool for limiting transmission. It is imperative to improve our range of biomedical interventions that can reduce viral transmission and to formulate a public-health strategy that also relies on passive nonpharmaceutical interventions (such as improved ventilation and air quality). Widespread and systematic surveillance of viral transmission is also key to enable the rapid implementation of nonpharmaceutical interventions to limit the risk of sudden shifts in virulence.

As we grapple with the reality of a long-running pandemic, there is a strong temptation to cast current events in an optimistic light. The actual trajectory of the pandemic so far has been bleak beyond all projections-two years in, we now have over a million dead in the US alone, with rapidly waning vaccinal and natural immunity facing off against a virus that is much more contagious than the ancestral strain. Against this backdrop, the fact that the existing vaccines still work to prevent severe disease and death provides us a bulwark against catastrophe. The work presented here demonstrates the consequences if this last line of defense is breached by viral evolution. In doing so, it underscores a key reality for risk mitigation during this pandemic: that unthinkable and impossible are not the same thing. A greater focus needs to be placed on biomedical interventions and public health strategies that are robust to viral evolution.

## Methods

### Fitness disadvantage incurred by fatal patient outcomes

To calculate the viral fitness disadvantage incurred by COVID-19 fatalities, we estimated the fractional loss of transmission that occurs when a patient infected with SARS-CoV-2 dies. We assumed that the distribution of probability of transmission over time is independent of disease severity. We used previously published distributions describing the infectivity of COVID-19 patients over time ^42^ and the probability of fatal outcome over time during disease progression^43^. We implemented both distributions in Python to assess loss of transmissibility due to fatal COVID-19 outcomes. Transmissibility over time is represented by a gamma distribution, implemented in the Python Scipy stats module with shape 20.52, scale 1/1.59, and loc parameter −12.27 according to He et al ^42^. The likelihood of fatal outcome over time is represented by a log-logistic distribution in the Scipy stats module with scale 31.18, shape 6.80, and loc parameter –14.51 according to Bai et al ^43^. To determine the loss of transmission that occurs when a patient dies, we converted the probability distribution function (PDF) of time from symptom onset to death to a cumulative distribution function (CDF) representing likelihood of survival over time. Then, we performed the dot product of the transmissibility and survival distributions to determine probability of transmission before death, given that both events occur. The loss of transmission due to fatal outcome is 1 – the fractional transmissibility before fatal outcome.

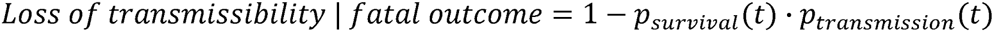

This loss of transmissibility occurs in the fraction of infections resulting in fatal outcome, which is the IFR. Thus, the overall loss of transmissibility is the IFR multiplied by the fractional loss of transmissibility in fatal cases.

### SEIRS modeling to predict endemic infection and death rates

To determine the impact of changes in SARS-CoV-2 properties R_0_, IFR, and duration of natural immunity on yearly US death tolls and infection rates, we varied these parameters in a susceptible-exposed-infectious-recovered-susceptible (SEIRS) epidemiological model under a series of vaccination conditions. The model contains two sets of SEIR compartments representing the differing infection and fatality rates of vaccinated and unvaccinated individuals. We assumed that the vaccine reduces the risk of death given infection (VEm) and the risk of infection (VEi) but has no additional impact on transmission in breakthrough cases. Some reports from earlier in the pandemic indicated a 50% reduction in infectiousness associated with vaccine breakthrough cases ^106^. However, recent contact-tracing findings, conducted in a household setting with omicron as the prevalent variant, showed no reduction in susceptibility to infection for breakthrough cases for fully-vaccinated individuals ^107^. Waning vaccinal effectiveness ^108^ in reducing viral load (which has now been noted for the booster dose as well ^109,110^ can be further expected to impact the vaccinal reduction of transmission. We ran the model under two theoretical vaccine acceptance scenarios in the US (70% uptake and 100% uptake). Optimistically, we assumed that the vaccine’s efficacy does not change over time (either due to high immunological durability or repeated boosting). Additionally, there is no age stratification in the model, so all parameter estimates represent averages across the US population.

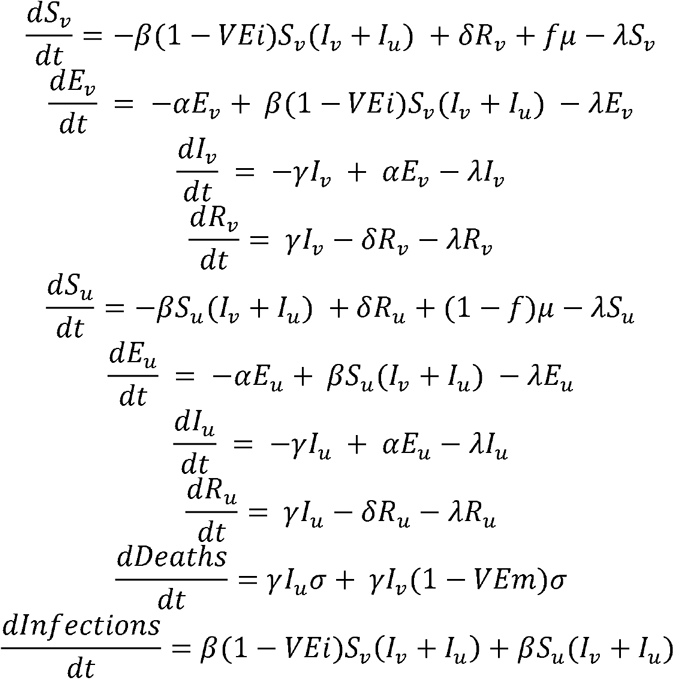

Subscripts *v* and *u* represent the vaccinated and unvaccinated subpopulations, respectively, in each of the SEIR model compartments. S represents susceptible individuals without immune protection; E represents exposed individuals who are not yet infectious; I represents infectious individuals; and R represents recovered individuals with natural immunity from infection. Cumulative deaths and infections are stored in separate variables (Deaths, Infections). The contact rate parameter *β* is a function of R_0_ according to *β* = *γR*_0_. In this analysis, we predicted yearly US infections while the duration of natural immunity and the R_0_ varied over a range. We also evaluated model-predicted yearly deaths over a range of IFRs and durations of immunity under multiple conditions for R_0_, vaccinal efficacy against infection, and vaccine uptake. The values of fixed model parameters are covered in Table 2, while the variable parameters and their ranges are covered in Table 3.

**Table 2:** Fixed parameters for SEIR model.

| Fixed Parameter | Symbol | Value | Source |
| --- | --- | --- | --- |
| Latency period | $1/\alpha$ | 3 days | <sup>111</sup> |
| Infectious period | $1/\gamma$ | 10 days | <sup>112</sup> |
| US population birth rate | $\mu$ | 0.9% annually | Fixed to death rate |
| US population death rate | $\lambda$ | 0.9% annually | <sup>113</sup> |
| US population size | N | 330 million | <sup>114</sup> |

**Table 3:** Variable parameters for SEIR model.

| Variable Parameter | Symbol | Value | Source |
| --- | --- | --- | --- |
| Vaccine reduction in risk of infection | VEi | 0%, 50%, 90% | <sup>115</sup> |
| Vaccine reduction in risk of mortality | VE<sub>m</sub> | 70%, 90% | <sup>116</sup> |
| Duration of natural immunity | $1/\delta$ | 3 to 24 months | <sup>117</sup> |
| Intrinsic reproductive number | $R_0$ | 2 to 9 individuals | <sup>118</sup> |
| IFR for unvaccinated individuals | $\sigma$ | 0.05% to 5% | See table S3 |
| Fraction vaccinated | f | 70%, 100% | <sup>90</sup> |

We used this model to estimate yearly US COVID-19 fatalities under endemic conditions, defined here as occurring when steady-state levels of disease spread are reached and maintained because the level of population immunity is equal to the herd immunity threshold (R_0_ – 1)/ R_0_. We assumed that all parameters are fixed within a single simulation (e.g., there are no time-dependent changes in any model parameters), and we ran the simulation for a long interval to ensure steady-state conditions were reached. We also note that in some high IFR scenarios, no steady state would be reached because deaths outpace new births, resulting in population loss. Given that we have demonstrated that COVID-19 mortality has very little impact on transmission, we neglected the impact of COVID-19 fatalities on population size – that is, we did not subtract fatal COVID-19 cases from the infected compartment. We also assumed the birth rate is equal to the death rate, so the population neither grows nor shrinks. This allows direct comparison of time-independent annual infection counts and death tolls between different IFR, vaccination, and immunity scenarios. We make the simplifying assumption that newborns are vaccinated or unvaccinated based on the overall fraction vaccinated.

## Data Availability

All data produced in the present study are available upon reasonable request to the author. Relevant code is available at https://github.com/madistod/endemicity

## Code availability

All simulations and analyses were implemented in Python, and code for running these simulations and plotting the results are available on GitHub.

## Supplementary Figures and Tables

**Figure S1.**
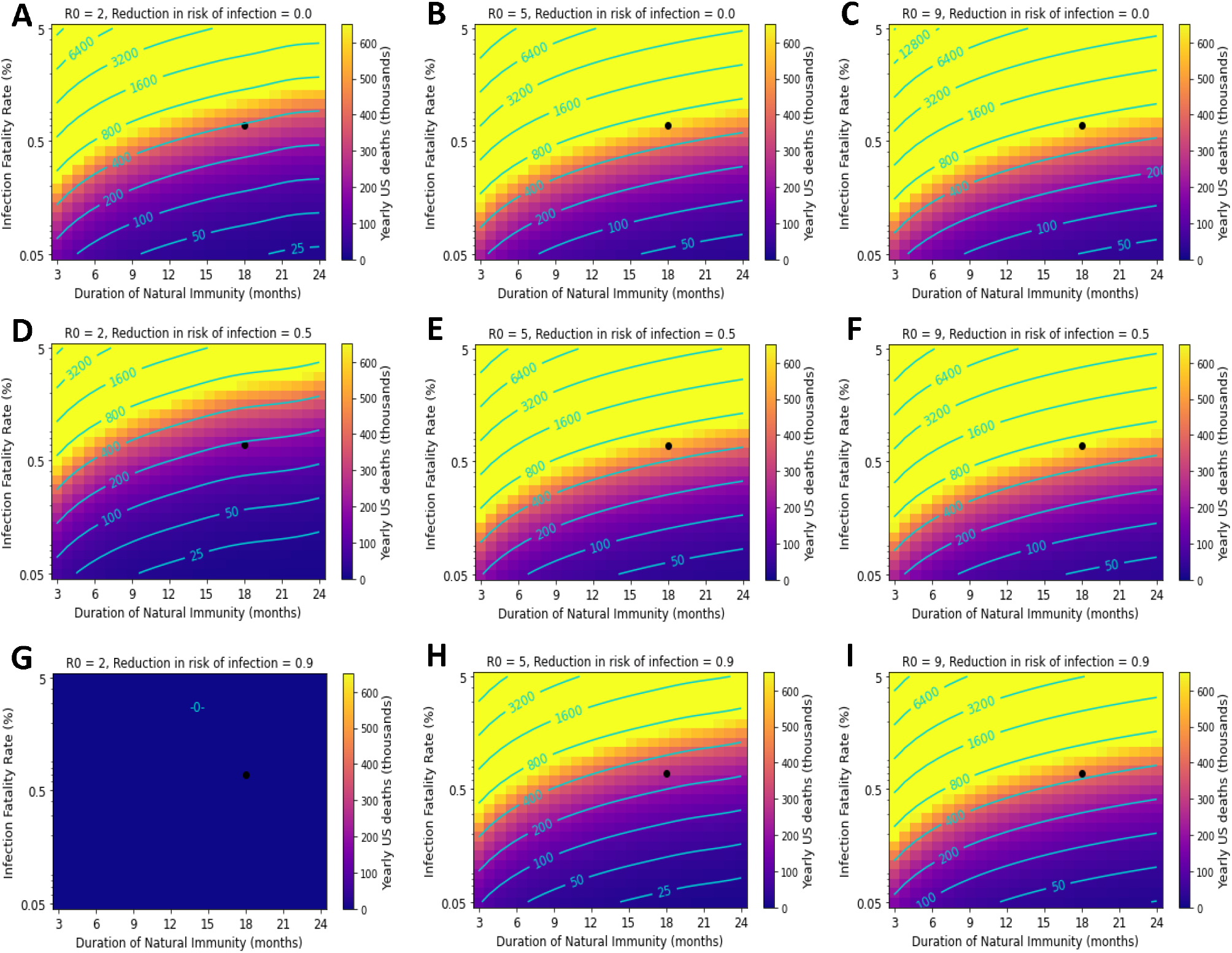
Death tolls are increased under poor vaccine performance. This figure mirrors Figure 3 but reduces VEm from 90% to 70%. Black point represents parameter values corresponding to best-estimates of immunity and IFR for ancestral SARS-CoV-2. Yearly US COVID-19 deaths under the following transmissibility (R_0_) and vaccine efficacy against transmission conditions: A-C) 0% VEi and R_0_ of 2, 5, and 9; D-F) 50% VEi and R_0_ of 2, 5, and 9. G-I) 90% VEi and R_0_ of 2, 5, and 9. Vaccine compliance is 70%.

**Figure S2.**
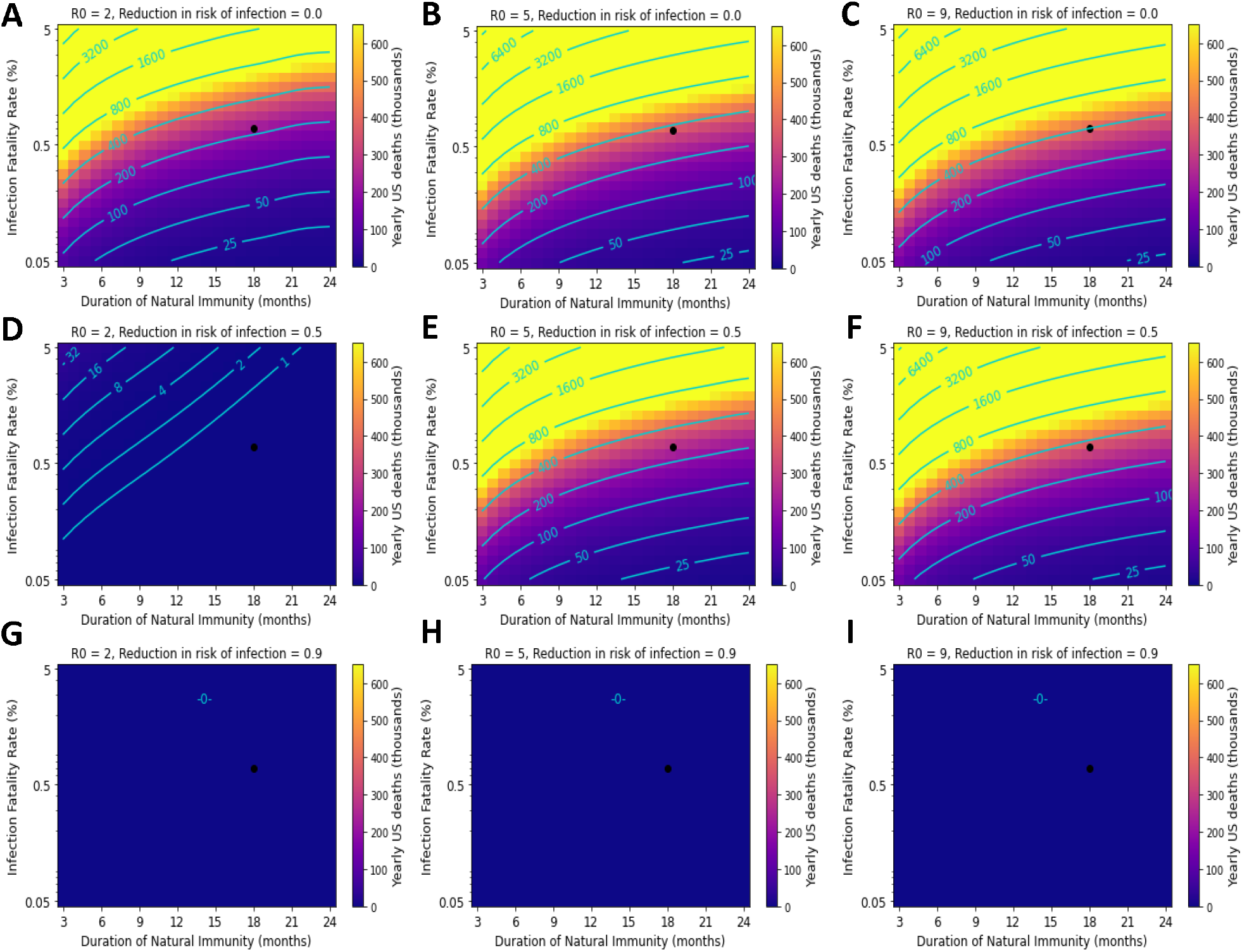
Suppression of SARS-CoV-2 transmission mitigates reduced vaccine effectiveness against mortality. This figure mirrors Figure 4 but reduces vaccine VEm from 90% to 70%. Black point represents parameter values corresponding to best-estimates of immunity and IFR for ancestral SARS-CoV-2. Yearly US COVID-19 deaths under the following transmissibility (R_0_) and vaccine efficacy against transmission conditions: A-C) 0% VEi and R_0_ of 2, 5, and 9; D-F) 50% VEi and R_0_ of 2, 5, and 9; G-I) 90% VEi and R_0_ of 2, 5, and 9. Vaccine compliance is 100%.

**Table S1.**
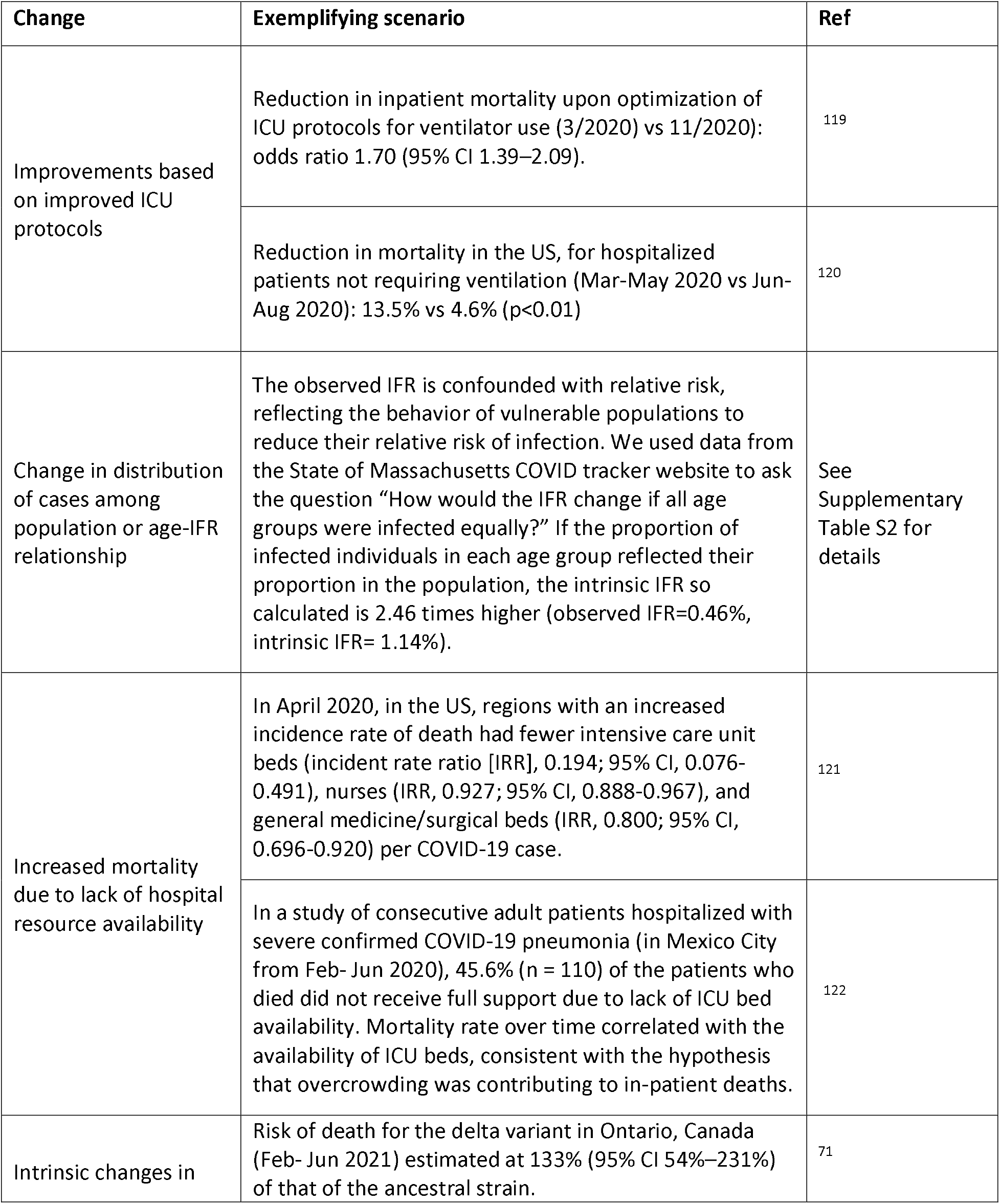
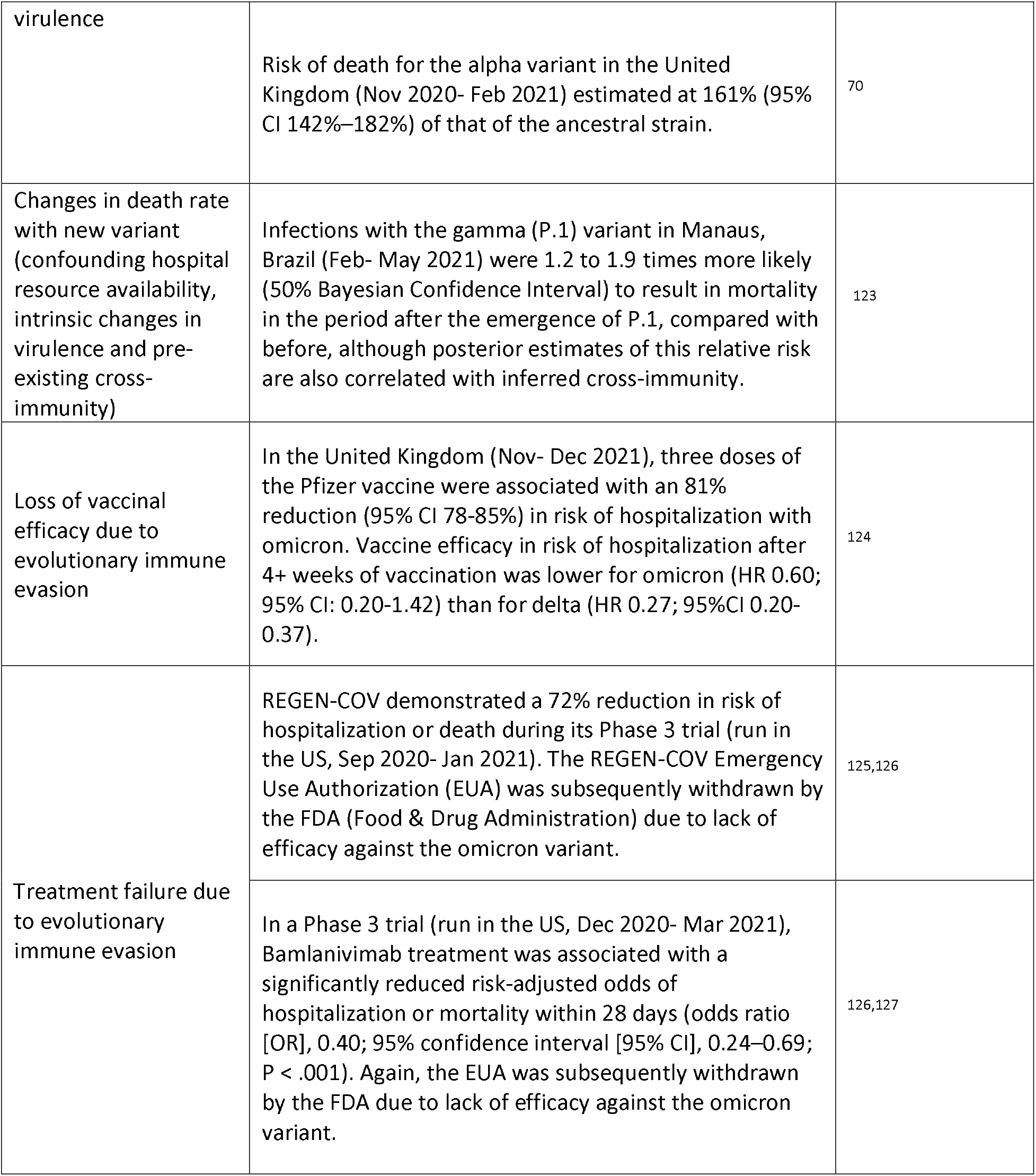
Illustrative examples of the changes observed so far in the IFR and risk of hospitalization during the pandemic (Mar 2020-Jan 2021):

**Table S2:** Estimation of the impact of shielding of older populations on apparent IFR using MA Covid tracker dataset (01/2021) as an example ^128,129^. In this analysis, we used published data on age-dependent COVID-19 IFRs to calculate the Massachusetts population average intrinsic IFR based on the MA population age structure. The apparent MA IFR deviates from this intrinsic IFR because infections are not distributed equally by age group, with older groups having lower infection rates. We determined the apparent MA population IFR as an average of the age-dependent IFRs weighted by age-dependent case rates.

| <u>MA Age group</u> | <u>IFR (*)</u> | <u>%pop (1)</u> | <u>%cases (1)</u> | <u>relative risk</u> | <u>Observed IFR</u> | <u>Contrib. Obs. IFR</u> | <u>Contrib. Int. IFR</u> | <u>Cases (1)</u> |
| --- | --- | --- | --- | --- | --- | --- | --- | --- |
| 0 to 4 | 0.0007 | 5.15% | 5.6% | 109% | 0.0007 | 3.8E-05 | 3.5E-05 | 8,923 |
| 5 to 9 | 0.0012 | 5.30% | 5.2% | 98% | 0.0012 | 6.5E-05 | 6.6E-05 | 8,268 |
| 10 to 14 | 0.0023 | 5.72% | 5.9% | 103% | 0.0024 | 0.00013 | 0.00013 | 9,388 |
| 15 to 19 | 0.0042 | 6.63% | 7.7% | 116% | 0.0048 | 0.00032 | 0.00028 | 12,232 |
| 20 to 29 | 0.0103 | 14.74% | 22.4% | 152% | 0.0157 | 0.00232 | 0.00152 | 35,740 |
| 30 to 39 | 0.0345 | 13.13% | 18.5% | 141% | 0.0486 | 0.00638 | 0.00453 | 29,453 |
| 40 to 49 | 0.1153 | 12.08% | 13.0% | 108% | 0.1242 | 0.01500 | 0.01393 | 20,726 |
| 50 to 59 | 0.3853 | 13.73% | 11.4% | 83% | 0.3199 | 0.04393 | 0.05290 | 18,160 |
| 60 to 69 | 1.2877 | 12.08% | 6.3% | 52% | 0.6707 | 0.08102 | 0.15555 | 10,023 |
| 70 to 79 | 4.3033 | 7.20% | 2.6% | 36% | 1.5399 | 0.11087 | 0.30984 | 4,104 |
| 80 + | 14.3814 | 4.21% | 1.4% | 34% | 4.8745 | 0.20522 | 0.60546 | 2,273 |

(1): data taken from MA covid tracker.
(2): regression formula from previously published meta-analysis<sup>128</sup> used to calculate IFR.
IFR= infection fatality rate
%pop= percentage of population
Contrib. Obs IFR= contribution of age group to observed IFR
Contrib. Int. IFR= contribution of age group to intrinsic IFR
**Observed IFR** 0.47
**Intrinsic IFR** 1.14
**Hazard Ratio (intrinsic/true)** 2.46

**Table S3:** IFRs and relative transmissibilities of SARS-CoV-2 ancestral strain and VoCs.

| <u>Variant</u> | <u>Infection fatality rate</u> | <u>Calculation</u> | <u>Transmissibility relative to ancestral strain</u> |
| --- | --- | --- | --- |
| Ancestral | 0.68% | <sup>46</sup> | 1.00 |
| Alpha | 1.09% | $1.6 \times 0.68\%$ <sup>70 **</sup> | $1.59$ <sup>45</sup> |
| Beta | 1.71% | $1.57 \times 1.6 \times 0.68\%$ <sup>130 ***</sup> | $1.50$ <sup>131</sup> |
| Gamma | 1.03% | $1.51 \times 0.68\%$ <sup>71 †</sup> | $2.00$ <sup>123</sup> |
| Delta | 1.58% | $2.33 \times 0.68\%$ <sup>71 ††</sup> | $1.82$ <sup>132</sup> |
| Omicron | 0.21% | $0.13 \times 2.33 \times 0.68\%$ <sup>133 †††</sup> | $2.13$ (est<sup>*</sup>) <sup>107</sup> |

\* $R_0$ of omicron is unknown, cited study found a secondary attack rate for household transmission between unvaccinated individuals that was 1.17-fold higher than delta. (2.6-3.6 fold higher than delta, in the vaccinated population).
\*\* estimated as 60% higher than the ancestral strain in the cited publication.
\*\*\* estimated as 57% higher than the alpha variant in the cited publication.
† estimated as 51% higher than the ancestral strain in the cited publication.
†† estimated as 133% higher than the ancestral strain in the cited publication.
††† estimated as 87% lower than the delta variant in the cited publication.

A. N. acknowledges funding from the National Science Foundation Graduate Research Fellowship Program under Grant No. DGE-1762114. Any opinions, findings, and conclusions or recommendations expressed in this material are those of the author(s) and do not necessarily reflect the views of the National Science Foundation.

